# Distinct pulmonary transcriptomic and proteomic host signatures enable accurate diagnosis of severe lower respiratory tract infection

**DOI:** 10.64898/2026.11.23.26368772

**Authors:** Emily C. Lydon, Peter Sax, Hoang Van Phan, Christina Osborne, Aline B. Maddux, Matthew K. Leroue, Nadir Yehya, BREATH Consortium, Lauren Gunn-Sandell, Matt S. Zinter, Joseph L. DeRisi, Brandie Wagner, Lilliam Ambroggio, Peter M. Mourani, Charles R. Langelier

## Abstract

Lower respiratory tract infection (LRTI) is a leading cause of acute respiratory illness, yet distinguishing it from noninfectious conditions remains a clinical challenge. Measuring the host immune response in the blood or lower airways, rather than exclusively detecting pathogens, has emerged as an alternative diagnostic strategy. However, it remains unclear whether RNA or protein measurements yield greater diagnostic information, or whether the two capture the same underlying biology. To address this, we performed paired transcriptomics and proteomics on tracheal aspirate specimens from 105 critically ill children with respiratory failure, 72 with microbiologically confirmed LRTI and 33 with noninfectious causes. Both approaches detected a clear host signature of LRTI which converged on similar core immune pathways including interferon signaling, inflammation, and complement activation. Despite this, agreement between individual genes and their corresponding proteins was largely poor. Diagnostic classifiers built from either RNA or protein data identified LRTI with similarly high accuracy (AUC>0.90), but selected different top biomarkers, and yielded discordant classifications for 12% of patients. Taken together, we find that lower airway transcriptomics and proteomics capture similar global biological features of LRTI despite poor correlation between gene and protein expression, providing fresh insight into approaches for biomarker discovery and clinical translation.

## INTRODUCTION

Lower respiratory tract infection (LRTI) is a leading cause of morbidity and mortality in children worldwide and a common indication for pediatric intensive care admission.^1–4^ In a child with undifferentiated respiratory failure, establishing whether LRTI is the underlying etiology is challenging, as noninfectious causes can present with overlapping clinical, radiographic, and laboratory features. Diagnosis primarily hinges on detecting a causative organism by culture, antigen testing, or nucleic acid amplification. However, such tests can be slow, detect only a narrow range of pathogens, have limited sensitivity, and be impacted by preceding antibiotics, leaving many cases of suspected LRTI without an etiologic diagnosis.^5–7^ Conversely, merely detecting a potential pathogen does not establish that it is causing disease, since children frequently carry respiratory pathogens without true infection.^8–10^ As a result, LRTI treatment is often empiric, contributing to antimicrobial overuse, resistance, and missed alternative diagnoses.^11–13^ Profiling the host response has emerged as a complementary diagnostic strategy.^14^ Rather than asking whether a pathogen is present, it asks whether the patient is eliciting a host immune response consistent with infection.

Host transcriptional profiling is the most established of these approaches. Blood gene expression signatures can distinguish bacterial from viral infection and infectious from noninfectious illness in both adults and children,^15–19^ and several have been translated onto rapid, targeted platforms suitable for clinical use.^16,20^ Far fewer studies have profiled the lower airway directly, despite it being the site of disease in LRTI. Our group has demonstrated that lower airway gene expression signatures accurately identify LRTI in critically ill adults and children, and additionally distinguish LRTI from incidental pathogen carriage.^21–23^

Proteins are more proximal effectors of the immune response compared to transcripts, and thus proteomic profiling in theory could capture additional infection-related inflammatory data missed by transcriptomics alone. Systems-level proteomic approaches are less established as infectious disease diagnostic discovery tools relative to transcriptomics, and work to date has been almost entirely blood-based. Serum protein signatures have been developed for active pulmonary tuberculosis,^24,25^ for discriminating the etiology of pneumonia in a malaria-endemic pediatric population,^26^ and for identifying bacterial infection in febrile infants.^27^ Airway proteomics, in contrast, is comparatively unexplored, limited to the respiratory mucosal proteome in influenza^28^ and a recent proteomic profiling study of tracheal aspirate (TA) in pediatric viral LRTI.^29^ Respiratory proteomics has not yet been evaluated as a modality for the diagnosis of all-cause LRTI.

Because transcripts and translated proteins both originate from the same genes, RNA and protein measurements are often treated as interchangeable proxies. However, quantitative studies argue otherwise. Across studies spanning human samples and cell lines, correlation between mRNA and protein levels is generally modest and highly analyte-dependent.^30–33^ Multiple steps separate mRNA abundance from protein abundance, including translational efficiency, differences in mRNA and protein half-life, microRNA-mediated silencing, and regulation by RNA-binding proteins.^32,34,35^ Moreover, airway sampling may amplify this discordance, as RNA principally reflects transcription by resident and recruited airway cells, while protein in TA fluid may additionally reflect secreted mediators, plasma-derived proteins, and products of cell injury.^36,37^ Transcriptomics and proteomics may therefore provide useful and largely non-redundant readouts of the same system.^38^

Multi-omic studies of infection are increasingly common, and integrative frameworks have been applied to COVID-19 severity and recovery,^39,40^ to sepsis subphenotyping,^41^ and to host-pathogen biology more broadly.^42^ These studies are designed to combine molecular layers into a single model rather than compare them, and they generally do not report whether the layers agree. One small study by Sivakumaran et al. explored both proteomic and transcriptional profiling and developed classifiers to distinguish bacterial and viral LRTI, but this study did not assess gene-protein discordance, and host profiling was performed on peripheral blood.^43^ No study has performed paired transcriptomic and proteomic profiling of the same lower airway specimens in LRTI, leaving unresolved whether the two modalities capture the same host biology, nominate the same biomarkers, or identify the same patients as infected.

Here, we address this gap in a prospective cohort of critically ill children with acute respiratory failure requiring mechanical ventilation. We performed paired host transcriptomic and proteomic profiling on the same TA specimens from patients adjudicated as having either microbiologically confirmed LRTI or an alternative, noninfectious etiology of respiratory failure. We first compared differential expression and pathway-level enrichment between the two modalities to determine whether they implicate the same LRTI biology. We then evaluated concordance for individual gene-protein pairs and examined how each modality relates to pathogen burden. Finally, we compared multiple transcriptomic and proteomic classifiers, ranging from full regularized regression models to single-analyte tests, to assess whether the choice of measurement modality affects diagnostic performance and the classification of individual patients.

## RESULTS

### Patient cohort and clinical adjudication

Children with acute respiratory failure requiring mechanical ventilation for at least 72 hours were prospectively enrolled at eight hospitals in the Collaborative Pediatric Critical Care Research Network between February 2015 and December 2017.^44^ TA was collected within 24 hours of intubation, and host gene expression was profiled by RNA sequencing in 261 patients, as previously reported (**Figure S1**).^23^ These transcriptomic data formed the basis for the present study, in which we additionally performed proteomic profiling on a subset of the cohort with remaining TA.

LRTI status was adjudicated retrospectively by at least two study-site physicians with expertise in pediatric critical care and/or infectious diseases, who reviewed all clinical, laboratory, microbiologic, and radiographic data available through the end of admission, incorporated CDC/NHSN PNU1 criteria,^45^ and were blinded to sequencing results. Patients were assigned to one of four categories based on the combination of clinical diagnosis and standard-of-care microbiologic testing: “Definite LRTI” (clinical diagnosis of LRTI with supportive microbiology), “Suspected” (clinical diagnosis of LRTI without microbiologic findings), “Indeterminate” (microbiologic findings without clinical suspicion of LRTI), and “No Evidence” (a clear non-infectious cause of respiratory failure with neither clinical nor microbiologic evidence of LRTI).

Because the aim of this study was to compare host transcriptional and proteomic responses between unambiguous infection and non-infection states, proteomic profiling was subsequently performed on patients in the Definite LRTI and No Evidence groups with available residual TA (**Figure 1**). Of these, 105 had paired host gene expression and proteomic measurements and comprised the analytic cohort, including 72 patients with LRTI and 33 No LRTI controls.

**Figure 1.**
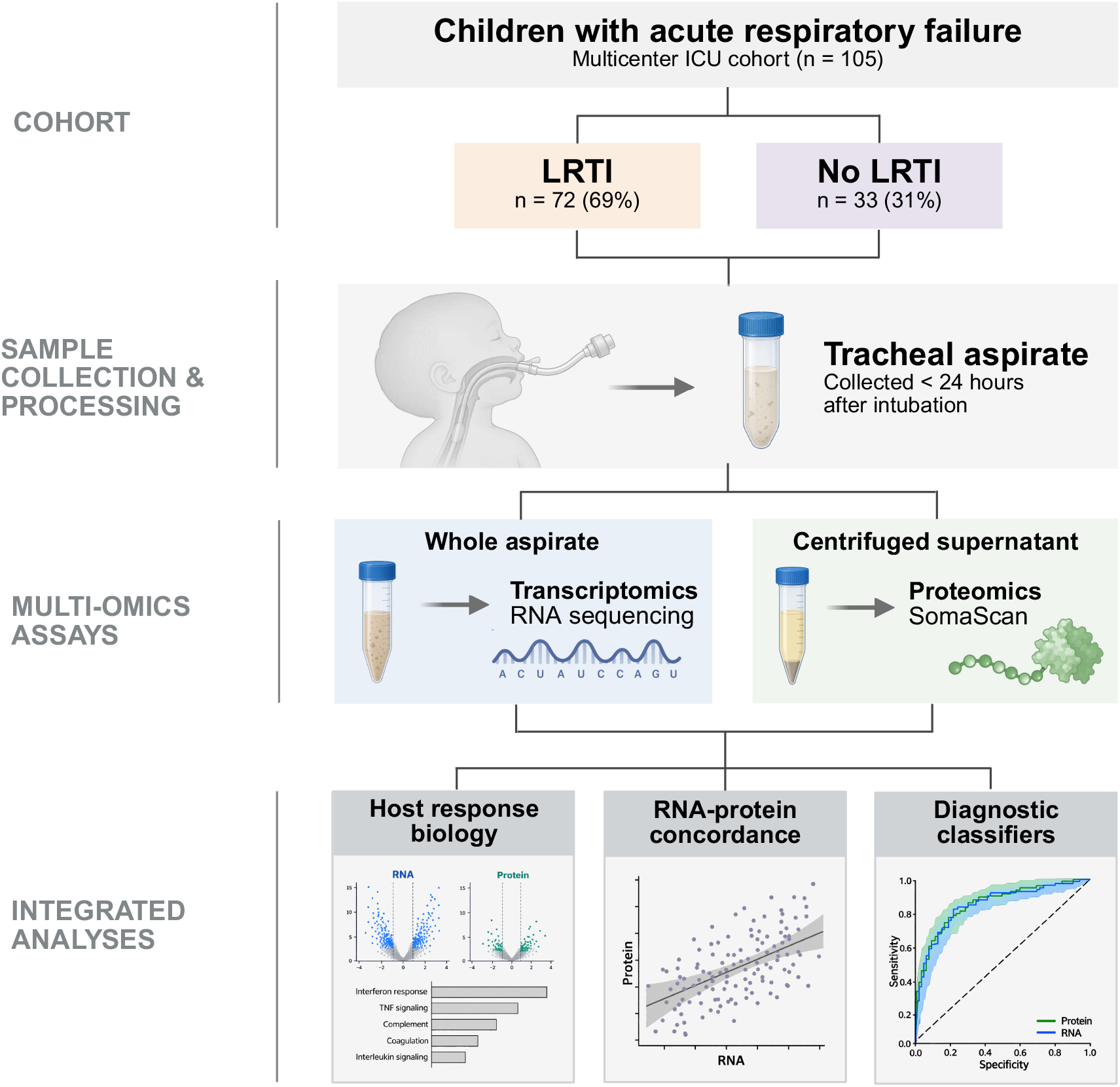
Study cohort and multi-omics analysis workflow. Children with acute respiratory failure enrolled in a multicenter ICU cohort were classified as having lower respiratory tract infection (LRTI; n = 72) or no LRTI (n = 33). Tracheal aspirates collected within 24 hours of intubation underwent host transcriptomic and proteomic profiling. Whole aspirate was analyzed by RNA sequencing, while centrifuged supernatant was analyzed by SomaScan proteomics. Integrated analyses evaluated host-response biology using differential expression and pathway enrichment, concordance between paired RNA and protein analytes, and diagnostic performance of transcriptomic and proteomic classifiers.

Demographic, clinical, and microbiologic characteristics of the analytic cohort are outlined in **Table 1**. Consistent with the epidemiology of pediatric respiratory failure and with prior analyses of this cohort,^21,23,46,47^ patients with LRTI were younger than controls (median 0.6 years, IQR 0.2-3.2 versus 5.3 years, IQR 1.3-11.3; P<0.001). Sex, race, ethnicity, and comorbidity burden did not differ significantly between groups. Admission category differed, with 100% of LRTI patients admitted for medical indications compared with 51.5% of controls, who more frequently had surgical or trauma admissions (P<0.001). Ventilator days, intensive care unit (ICU) length of stay, and in-hospital mortality were similar between groups, while hospital length of stay was modestly shorter in the LRTI group (16.0 [IQR 11.0-24.2] vs 19.0 [14.0-43.0] days, P=0.048). Antibiotic exposure prior to admission was similar between groups (40.3% versus 30.3%), and nearly all patients received antibiotics during hospitalization (100% versus 87.9%, P=0.009). Among patients with LRTI, clinical microbiologic testing identified a viral pathogen alone in 29 (40.3%), a bacterial pathogen alone in 13 (18.1%), and both in 30 (41.7%). The most commonly detected viral pathogens were respiratory syncytial virus (61.1%), non-COVID-19 coronavirus (9.7%), and adenovirus (6.9%), and the most commonly detected bacterial pathogens were *Moraxella catarrhalis* (22.2%), *Haemophilus influenzae* (19.4%), and *Staphylococcus aureus* (15.3%) (**Figure S2**).

**Table 1.** Demographic and clinical characteristics of patients with and without lower respiratory tract infection (LRTI). Race is indicated as “unknown” if patient declined or was unable to answer, if they selected “other” as an option, or if data were missing. Continuous variables are reported as median (interquartile range, IQR) and compared using the Wilcoxon rank-sum test. Categorical variables are reported as n (%) and compared using Fisher’s exact test. ICU, intensive care unit.

|  | **LRTI**<br>**(n = 72)** | **No LRTI**<br>**(n = 33)** | **P value** |
|---|---|---|---|
| **Female, n (%)** | 26 (36.1) | 17 (51.5) | 0.20 |
| **Male, n (%)** | 46 (63.9) | 16 (48.5) | 0.20 |
| **Age in years, median (IQR)** | 0.6 (0.2–3.2) | 5.3 (1.3–11.3) | <0.001 |
| **Race, n (%)** |  |  | 0.25 |
| **White** | 41 (56.9) | 21 (63.6) |  |
| **Black or African American** | 18 (25.0) | 3 (9.1) |  |
| **Asian** | 3 (4.2) | 4 (12.1) |  |
| **Native Hawaiian/Pacific Islander** | 1 (1.4) | 0 (0.0) |  |
| **Multi-racial** | 3 (4.2) | 1 (3.0) |  |
| **Unknown** | 6 (8.3) | 4 (12.1) |  |
| **Hispanic/Latino ethnicity, n (%)** | 8 (11.1) | 5 (15.2) | 0.54 |
| **Chronic medical condition, n (%)** | 31 (43.1) | 18 (54.5) | 0.30 |
| **Admission category, n (%)** |  |  | <0.001 |
| **Medical** | 72 (100.0) | 17 (51.5) |  |
| **Surgical** | 0 (0.0) | 11 (33.3) |  |
| **Trauma** | 0 (0.0) | 5 (15.2) |  |
| **Antibiotics prior to admission, n (%)** | 29 (40.3) | 10 (30.3) | 0.39 |
| **Any antibiotic use, n (%)** | 72 (100.0) | 29 (87.9) | 0.009 |
| **Systemic corticosteroids, n (%)** | 18 (25.0) | 5 (15.2) | 0.32 |
| **Ventilator days, median (IQR)** | 7.0 (4.8–9.0) | 5.0 (4.0–8.0) | 0.37 |
| **ICU length of stay, days, median (IQR)** | 10.5 (7.0–15.2) | 9.0 (7.0–15.0) | 0.99 |
| **Hospital length of stay, days, median (IQR)** | 16.0 (11.0–24.2) | 19.0 (14.0–43.0) | 0.048 |
| **In-hospital mortality, n (%)** | 1 (1.4) | 2 (6.1) | 0.23 |
| **Microbiologic diagnosis, n (%)** |  |  |  |
| **Viral only** | 29 (40.3) |  |  |
| **Bacterial only** | 13 (18.1) |  |  |
| **Viral/bacterial coinfection** | 30 (41.7) |  |  |

### LRTI is associated with broad transcriptomic and proteomic changes that converge at the pathway level

We first asked whether LRTI produced a detectable host signature in each modality. Differential expression analysis identified 1,034 differentially expressed genes between LRTI and control patients (7.4% of 13,941 genes tested at P_adj_<0.05; **Figure 2A**), while differential abundance analysis identified 40 differentially abundant proteins (0.6% of 6,346 proteins measured at P_adj_<0.05; **Figure 2B**). Both modalities were characterized by canonical antiviral and inflammatory mediators. Interferon-stimulated genes and proteins including ISG15, IFIT3, and MX1 were among the most strongly upregulated in LRTI,^48^ as were the chemokine CCL8 and the cytotoxic effectors GNLY and GZMB.^49^ FABP4, an inflammatory mediator and marker of alveolar macrophages, was the most significantly downregulated analyte in both transcriptome and proteome in LRTI.^50,51^

**Figure 2.**
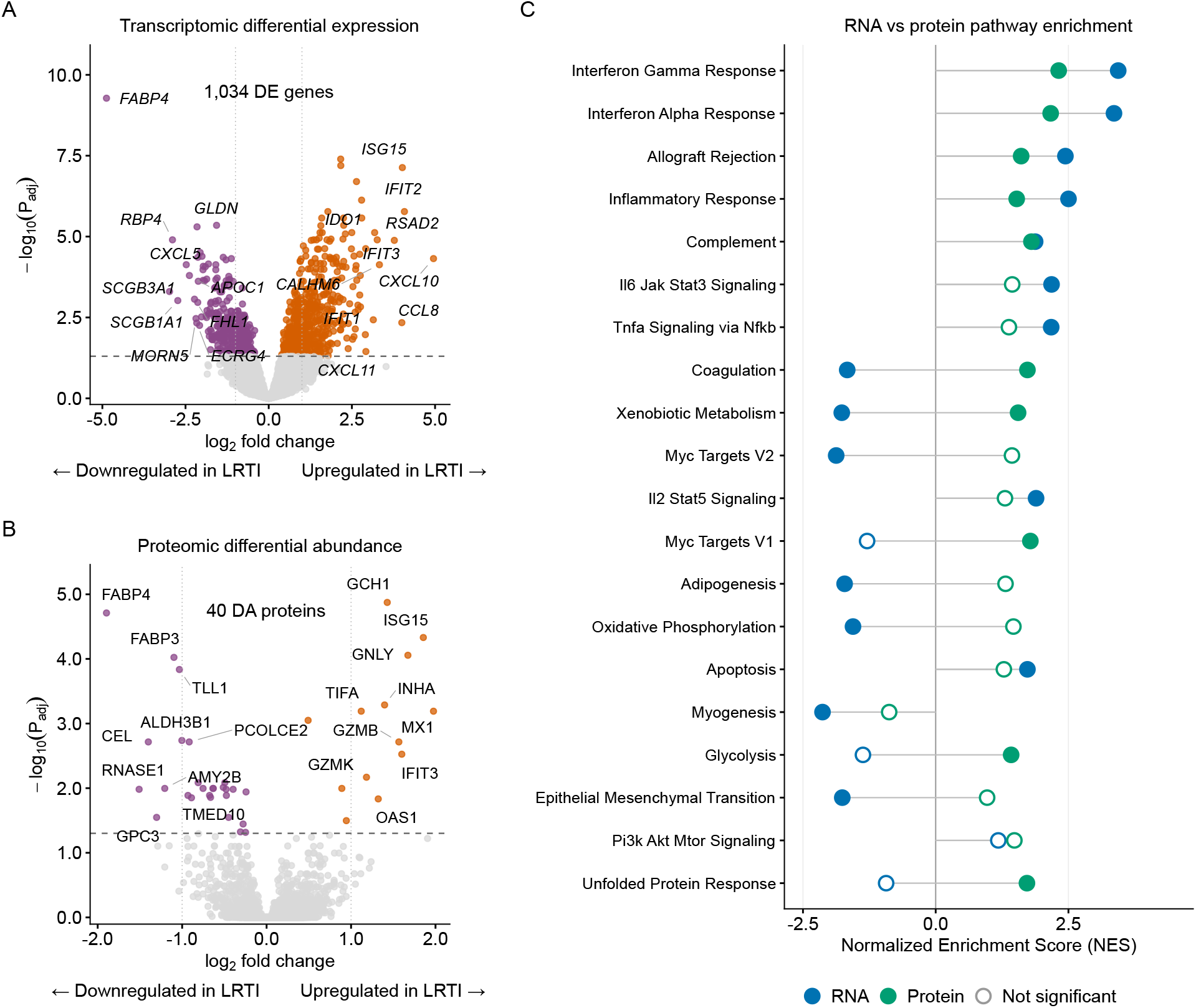
LRTI is associated with broad transcriptomic and proteomic changes in the lower airway that converge at the pathway level. A. Volcano plot of differential gene expression in tracheal aspirate samples between patients adjudicated as LRTI (n=72) and those with no evidence of LRTI (n=33), measured by limma-voom and adjusted for age and sex. (B) Volcano plot of differential protein abundance between the same groups, based on SomaScan proteomic profiling tracheal aspirate. In both panels, the x-axis shows log_2_ fold change and the y-axis shows −log_10_ of the Benjamini-Hochberg adjusted P value. Analytes significantly upregulated in LRTI are shown in orange, those significantly downregulated in purple, and non-significant in grey. C. Gene set enrichment analysis of MSigDB Hallmark gene sets, performed separately on the transcriptomic (blue) and proteomic (green) differential expression/abundance results ranked by log_2_ fold change. The 20 pathways most significantly enriched in the transcriptomic analysis are shown. Points indicate the normalized enrichment score (NES) for each modality. Filled points denote statistically significant enrichment (P_adj_ < 0.05) and open points denote non-significant enrichment. Positive NES indicates enrichment in LRTI. LRTI, lower respiratory tract infection; DE, differentially expressed; DA, differentially abundant.

To compare the two modalities at a broader pathway level, we performed gene set enrichment analysis on each dataset independently and compared normalized enrichment scores (**Figure 2C**). Pathways canonically associated with infection were concordantly enriched in LRTI in both modalities, including interferon-α and interferon-γ responses, inflammatory responses, complement, IL-6/JAK/STAT3 signaling, and TNF-α signaling via NF-κB. Several pathways, however, demonstrated enrichment scores in opposite directions; coagulation and xenobiotic metabolism were significantly depleted in the transcriptome but significantly enriched in the proteome. These discordances may reflect differences in what each modality captures, such as accumulation of plasma-derived coagulation proteins in the injured airway independent of local transcription.^52,53^ Nonetheless, the two modalities recovered a broadly convergent picture of the host response to LRTI at the pathway level.

### RNA and protein measurements demonstrate limited analyte-level concordance

Having found that both modalities recovered a largely convergent pathway-level picture of the host response, we next asked whether this agreement extended to individual analytes. We identified 6,238 analytes measured by both platforms and computed the Pearson correlation between RNA and protein levels across all 105 patients. Correlations were centered near zero (median r = 0.021; IQR -0.06 - +0.11) (**Figure 3A**), and a minority of analytes showed appreciable concordance, with only 44 (0.71%) exceeding r = 0.5.

**Figure 3.**
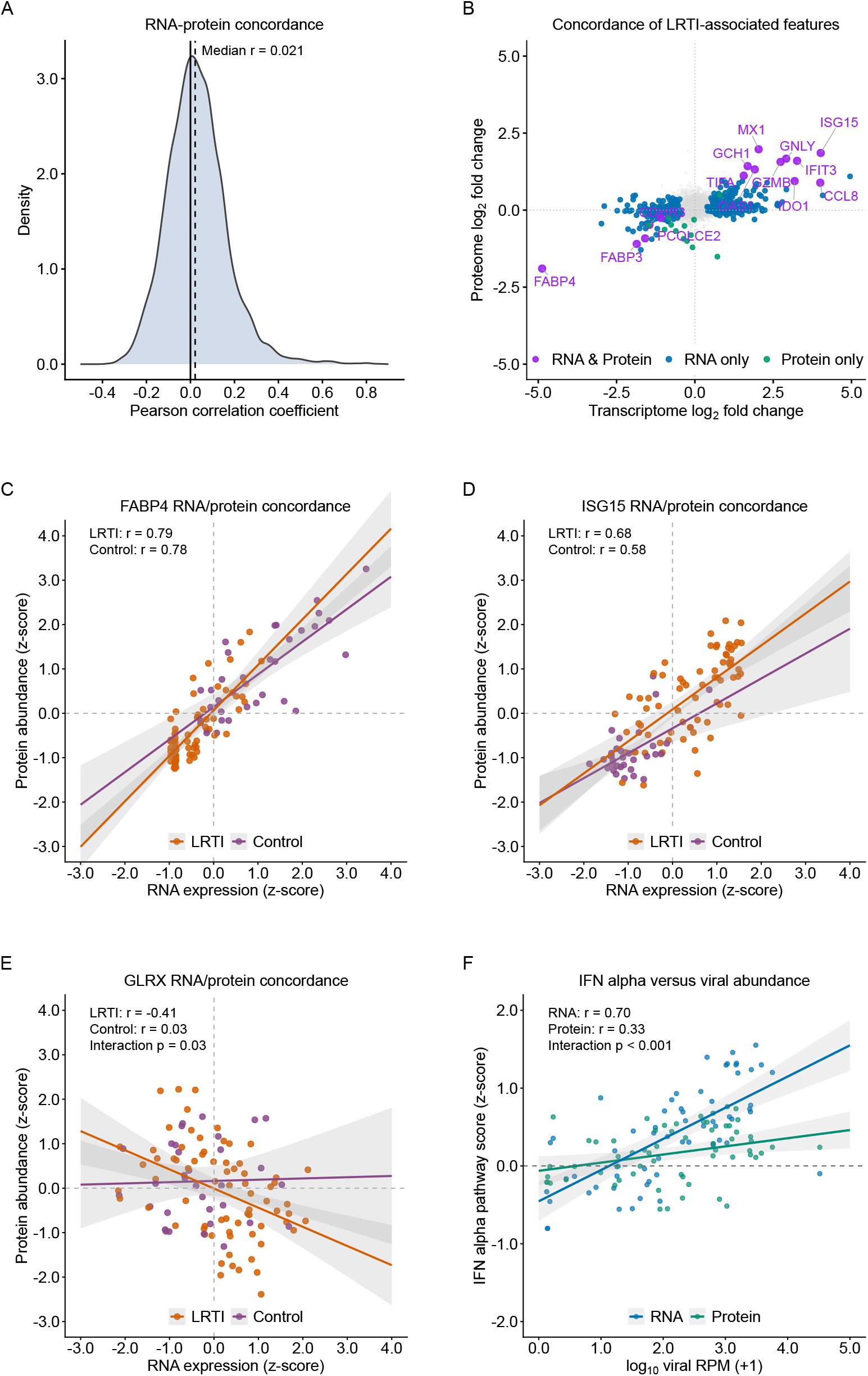
RNA and protein measurements of the same analytes show limited concordance at the individual analyte level. A. Density distribution of Pearson correlation coefficients between paired RNA and protein measurements for the analytes quantified on both platforms, calculated across all 105 patients using z-scored values. The dashed vertical line indicates the median correlation (r=0.021). B. Transcriptomic log_2_ fold change plotted against proteomic log_2_ fold change for each shared analyte, comparing LRTI patients (n=72) with controls (n=33). Analytes significantly differentially expressed by limma-voom in both modalities are shown in purple, transcriptome only in blue, proteome only in green, and not significant in either modality in grey. Selected concordant analytes are labeled. Benjamini-Hochberg adjusted P value < 0.05 utilized for significance. C–E. Scatterplots of z-scored protein abundance against z-scored RNA expression for FABP4 (C), ISG15 (D), and GLRX (E), stratified by LRTI status (orange = LRTI, purple = controls). Points represent individual patients, lines indicate linear model fits, and shaded bands indicate 95% confidence intervals. Within-group Pearson correlation coefficients are annotated, and differences in slope between groups were tested using a linear model with an RNA-by-group interaction term, which is shown if significant. F. Composite Hallmark “interferon alpha response” score, calculated separately for the transcriptome (blue) and proteome (green) as the mean of z-scored values across analytes in the gene set, plotted against log_10_ transformed summed viral abundance (reads per million, RPM) among LRTI patients with a respiratory virus detected by metatranscriptomic sequencing (n=62). Lines indicate linear model fits with 95% confidence intervals. Pearson correlation coefficients and the P value for the modality-by-viral-abundance interaction term are annotated. LRTI, lower respiratory tract infection.

To determine whether the analytes associated with LRTI were shared across modalities, we compared transcriptomic and proteomic fold changes for each analyte measured by both platforms (**Figure 3B**). Of the shared analytes, 15 were significantly differentially expressed in both the transcriptome and the proteome, all in the same direction; these included ISG15, MX1, GNLY, IFIT3, GZMB, CCL8, and FABP4. A larger number were significant in one modality only (357 in RNA alone, 13 in protein alone); the vast majority were not significant in either. No analyte was significantly differentially expressed in opposite directions.

We next examined whether the analytes most strongly associated with LRTI also exhibited the highest correlation between RNA and protein expression, and whether the correlation differed by LRTI status. FABP4 and ISG15 transcript and protein levels were strongly and positively correlated in both LRTI and controls (FABP4: r=0.79 in LRTI, r=0.78 in controls; ISG15: r=0.68 and r=0.58, respectively; **Figure 3C-D**), indicating that RNA-protein coupling for these analytes was preserved irrespective of infection status. For other analytes, however, the relationship between RNA and protein markedly differed based on presence or absence of LRTI. For instance, GLRX transcript and protein levels were not correlated in control patients (r = 0.03) but inversely correlated in patients with LRTI (r=-0.41; interaction P=0.032; **Figure 3E**). GLRX encodes glutaredoxin-1, an oxidative stress–responsive enzyme that regulates reversible protein S-glutathionylation, a key form of post-translational protein regulation.^54,55^ State-dependent patterns were also observed for SLAMF7, an immunoregulatory receptor expressed on multiple immune-cell populations (r=0.16 for LRTI, r=-0.38 for control, interaction P=0.004),^56^ and FAH, the terminal enzyme in tyrosine catabolism (r=-0.44 for LRTI, r=0.00 for control, interaction P=0.01) (**Figure S3**).^57^ Together, these findings suggest that RNA-protein coupling can be analyte-specific and differ based on presence or absence of infection in the lower airways.

### Pathogen burden associates with host response differently across modalities

We next asked whether the relationship between pathogen burden and host response differed based on assessment at the transcript or protein level. Among LRTI patients with a virus detected (n=62), we compared summed viral abundance (in metatranscriptomic reads-per-million) with a composite interferon alpha score calculated separately from transcriptomic and proteomic data (**Figure 3F**). Both modalities showed a positive association with viral load, consistent with prior literature,^58,59^ but the relationship was substantially stronger at the RNA versus protein level (r=0.70 vs r=0.33, respectively), and also characterized by a greater induction in analyte expression per given amount of virus (interaction P < 0.001). This difference persisted when the interferon score was restricted to analytes measured on both platforms (n=49 analytes; RNA r=0.68, protein r=0.33, interaction P< 0.001; **Figure S4**), indicating that it was not simply explained by differences in the analytes available to each assay.

We performed an analogous analysis of bacterial burden in the lower airway versus host response among LRTI patients with a bacterial pathogen detected (n=43), examining canonical antibacterial response analytes (CD177, MMP8, TLR2, TLR4; **Figure S5**) and neutrophil degranulation pathway scores (**Figure S6**). In contrast to the viral analysis, we observed no consistent association between bacterial pathogen abundance and host response in either modality. This may reflect a more complex relationship between bacterial abundance and infection in the lower airway, and whether transcriptomic and proteomic measurements diverge consistently in their relationship to bacterial pathogen burden remains unclear.

### Transcriptomic and proteomic classifiers provide comparable but non-identical diagnostic information

Given that the two modalities agreed at the pathway level but diverged for most individual analytes, it remained unclear whether transcriptomic and proteomic classifiers for LRTI diagnosis would perform comparably or capture the same diagnostic information. We thus trained random forest classifiers separately on transcriptomic and proteomic data using 5-fold cross-validation, with features selected by LASSO logistic regression within training folds at two levels of sparsity: a full model (lambda.1se) and a parsimonious model constrained to three features per fold (**Tables S1-S4**). We additionally evaluated single-analyte classifiers based on FABP4 and ISG15, selected for their prominence in the preceding analyses and established utility as single gene biomarkers of all-cause and viral LRTI, respectively.^29,60^

All classifiers discriminated LRTI from control patients with high accuracy (**Figure 4A**). The full LASSO models performed best, with pooled area under curves (AUCs) of 0.93 (95% CI 0.87-0.98) for the transcriptome and 0.93 (0.87-0.99) for the proteome. Restricting to three features reduced performance only modestly (transcriptome 0.89 [0.82-0.96]; proteome 0.91 [0.84-0.97]; (**Figure 4B-C**), indicating that most of the diagnostic signal in each modality was captured by a small number of analytes. Performance did not differ significantly between transcriptomic and proteomic classifiers at either level of sparsity (DeLong P = 0.99 and P = 0.66).

**Figure 4.**
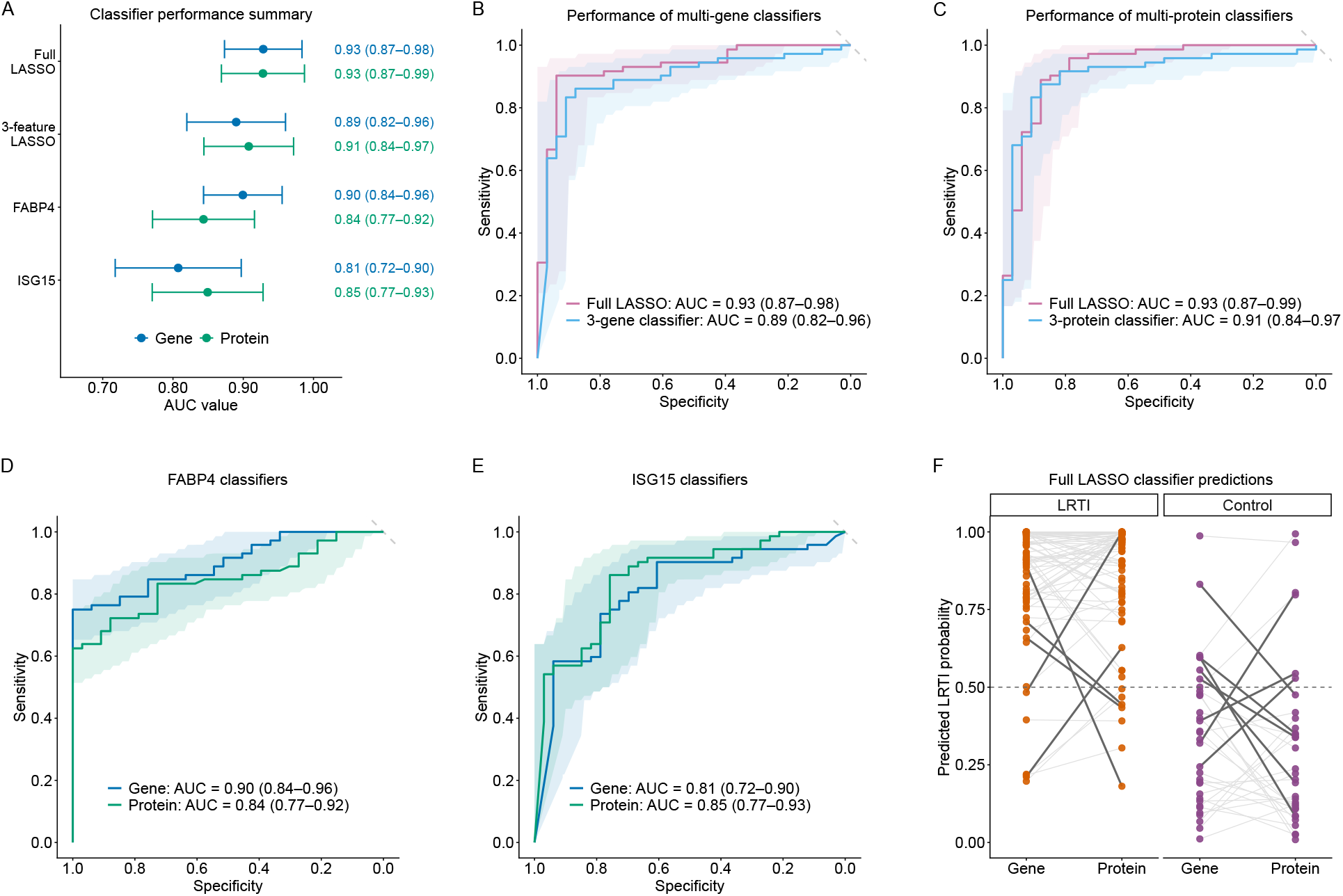
Transcriptomic and proteomic classifiers provide comparable but non-identical diagnostic information. A. Pooled area under the receiver operating characteristic curve (AUC) for each classifier, built on transcriptomic (blue) and proteomic (green) data. Points indicate the AUC and horizontal lines the 95% confidence interval; values are annotated at right. Classifiers were trained as random forests on features selected by LASSO logistic regression within training folds at two levels of sparsity (full model, lambda.1se; 3-feature model), alongside single-analyte classifiers FABP4 and ISG15. All classifiers were evaluated by stratified 5-fold cross-validation, and AUCs were computed from pooled out-of-fold predictions. B–C. Receiver operating characteristic curves comparing the full LASSO classifier (purple) with the 3-feature classifier (blue) for the transcriptome (B) and the proteome (C). D–E. Receiver operating characteristic curves comparing single-analyte classifiers measured as transcript versus protein for FABP4 (D) and ISG15 (E). In B–E, shaded bands indicate 95% confidence intervals, the diagonal dashed line indicates chance performance, and AUCs with 95% confidence intervals are annotated. F. Out-of-fold predicted probability of LRTI from the transcriptomic and proteomic full LASSO classifiers for each patient, faceted by adjudicated LRTI status (LRTI in orange, control in purple). Each line connects the two predictions for a single patient; dark lines indicate the 13 patients (12.4%) whose predictions crossed the 50% classification threshold between modalities. LRTI, lower respiratory tract infection.

Single-analyte classifiers performed well considering their simplicity, but the more informative modality differed by analyte. FABP4 achieved a higher AUC at the transcript versus protein level (AUC 0.90 [0.84-0.96] versus 0.84 [0.77-0.92]; **Figure 4D**), although the difference did not reach statistical significance (DeLong P = 0.085). ISG15 trended in the opposite direction (0.81 [0.72-0.90] for gene versus 0.85 [0.77-0.93] for protein; DeLong P = 0.45; **Figure 4E**).

Finally, we compared out-of-fold predictions from the transcriptomic and proteomic full LASSO classifiers for each patient (**Figure 4F**). Although predictions were broadly concordant, 13 patients (12.4%) crossed the 50% probability threshold between modalities, including 5 of 72 LRTI patients (6.9%) and 8 of 33 (24.2%) controls. Thus, despite similar overall performance, the two modalities did not classify identical sets of patients, suggesting that transcriptomic and proteomic measurements carry partially non-redundant diagnostic information.

## DISCUSSION

Host response profiling is an emerging and promising approach to LRTI diagnosis, but whether transcriptomic and proteomic measurements provide similar biological and diagnostic information has not been established. Here, in a prospective multicenter cohort of critically ill children, we performed the first paired host transcriptomic and proteomic profiling of TA specimens in LRTI.

Both modalities detected a robust host response signature that reflected canonical antiviral and inflammatory programs of infection. Concordance was far more limited below the pathway level, however, with most individual transcript–protein pairs demonstrating poor correlation in terms of lower airway expression. Despite these differences, diagnostic classifiers derived from either modality distinguished LRTI from noninfectious respiratory failure with comparably high accuracy. Taken together, these findings provide a new resource for both pulmonary diagnostic test development and future multi-omic immunoprofiling studies in the respiratory tract.

Both transcriptomic and proteomic profiling identified host signatures reflecting established innate immune responses to infection. Gene set enrichment performed independently on each data type identified concordant enrichment of interferon-α and interferon-γ responses, complement, IL-6/JAK/STAT3 signaling, and TNF-α signaling. This concordance extended to individual analytes within these programs, which were differentially expressed in the same direction regardless of data type, including the interferon-stimulated genes and proteins MX1, IFIT3, and OAS1, the interferon-γ effectors GCH1 and IDO1, the chemokine CCL8, and the cytotoxic effectors GNLY and GZMB. Among the relatively few analytes with tightly correlated expression between gene and protein, two of the most highly correlated were ISG15, reflecting type 1 interferon response,^61^ and FABP4, reflecting a shift in alveolar macrophage populations and modulation of inflammatory lipid signaling.^60,62^ Both were among the analytes most strongly associated with LRTI, and their correlation was unaffected by presence or absence of infection. These concordant biomolecules are largely produced by the resident and recruited cells of the airway, including macrophages, monocytes, neutrophils, and infiltrating lymphocytes.^63–65^ Shared cellular origin, with transcript and protein deriving from the same compartment, is one plausible explanation for their high correlation.

Concordance between gene and protein expression, however, was the exception, with measurements diverging far more often than they agreed. Correlation across analytes measured on both platforms was centered near zero (median r=0.021), with only a small minority exceeding r=0.5. The two modalities also differed markedly in the scale of the LRTI host response they detected, with 1,034 differentially expressed genes (7.4% of the transcriptome) compared with 40 differentially abundant proteins (0.6% of the measured proteome). For a subset of analytes, coupling depended on disease state: for GLRX and FAH, transcript and protein were unrelated in control patients but inversely correlated in patients with LRTI, whereas SLAMF7 showed the opposite pattern, inversely correlated at baseline and uncoupled during infection. Although most pathways were concordantly enriched between modalities, discordance was not limited to individual analytes: coagulation and xenobiotic metabolism were significantly enriched in opposite directions.

Several mechanisms may account for these discordances. RNA was sequenced from whole tracheal aspirate and therefore predominantly reflects transcription by the cells present, whereas protein was measured in the cell-free supernatant, where abundance may reflect a combination of local and systemic transcription.^36^ This distinction is most apparent for coagulation-related mediators, which is perhaps not surprising given that clotting cascades depend on hepatically synthesized plasma proteins that airway cells do not appreciably produce, and whose accumulation in the airway may reflect passage across an injured epithelial barrier rather than local production.^52,66^ Cell injury offers a distinct explanation for GLRX and FAH, intracellular proteins that are not normally secreted and therefore reach airway fluid largely through cell lysis.^67–69^ Dying cells may disproportionately release protein versus transcript, which could account for the inverse relationship between gene and protein expression for these biomolecules in the setting of LRTI. SLAMF7 illustrates a different potential mechanism for transcript-protein expression discordance. This cell surface receptor can be cleaved and released into surrounding fluid, so its protein abundance may reflect release from the cell surface rather than new transcription.^70^ Post-transcriptional and post-translational regulation likely contribute as well to transcript-protein discordance, as translation, protein stability, and turnover shape protein abundance independently of transcription.^32,71^ Finally, the kinetics of gene and protein expression differ; transcriptional changes can occur more rapidly than changes in protein accumulation or clearance, and a cross-sectional sample is intrinsically limited in terms of capturing this information.^30,72^ Consistent with this, the association between viral abundance and the interferon alpha response was substantially steeper in the transcriptome than the proteome, though no comparable relationship was observed for bacterial abundance.

Somewhat surprisingly, the limited correlation between gene and protein expression levels in the lower airway did not translate into a difference in diagnostic classifier performance. Classifiers derived from either modality performed comparably well, with full LASSO models achieving AUCs of 0.93 for both the transcriptome and the proteome, and sparser three-feature models performing nearly as well (AUCs of 0.89 and 0.91, respectively). With respect to single biomarkers, subtle differences in diagnostic performance were apparent. For instance, our data suggested that FABP4, the single best performing gene or protein biomarker for all-cause LRTI diagnosis, may perform more accurately as a transcript than as a protein. In contrast, the established viral infection biomarker ISG15 performed comparably in both modalities, if anything favoring protein. On the individual patient level, classifier prediction of infection status varied based on measurement platform. The transcriptomic and proteomic LASSO classifiers disagreed for 12.4% of patients, more often among controls than among those with LRTI, an asymmetry which may reflect the heterogeneity of noninfectious respiratory failure, in which a range of distinct processes produce host responses that the two modalities capture differently.

Beyond relatively small differences in classifier performance, practical considerations should also factor into how either modality might be translated into a diagnostic test. Sequencing offers broad coverage of the host response and can simultaneously capture pathogen information, and while costs continue to fall and access continues to broaden, it remains too complex and time consuming for direct clinical implementation in most clinical settings.^73,74^ Translation may more plausibly proceed through targeted quantification of a small number of transcripts by RT-qPCR, an approach already used in several blood-based host response assays now in development.^20,75,76^ Protein measurement may offer a more direct path to clinical deployment, as targeted immunoassays and lateral flow technology are established, inexpensive, and feasible without molecular diagnostic infrastructure, and several blood-based host protein signatures have already reached clinical implementation.^77,78^ In this context, the comparable performance of sparse classifiers in both modalities is encouraging, as a single- or three-analyte test is far more tractable than genome-wide measurement, and a modest performance cost may be acceptable if it buys a simpler, faster assay. Given limitations of pathogen-focused testing, progress toward a deployable host-based assay is needed,^14^ and our findings suggest either modality could support one.

This study has several strengths, including paired transcriptomic and proteomic profiling of the same specimens, sampling at the site of infection rather than in blood, and a prospectively enrolled multicenter cohort with rigorous case adjudication. Several limitations should also be considered. Our analyses were restricted to patients with clearly defined LRTI or noninfectious respiratory failure, which was appropriate for comparing biology across modalities and for classifier development, but excludes patients with uncertain infection status. Multi-physician retrospective clinical adjudication based on an established pneumonia definition is an imperfect gold standard, though it is the best available reference in the absence of a definitive diagnostic test. The aptamer-based platform used in this study, like most proteomics platforms, samples the proteome incompletely and may bias toward certain protein classes, and thus it is possible that the degree of transcript-protein discordance could vary based on platform used.^79–81^ Control patients were substantially older than patients with LRTI, and although differential expression analyses were adjusted for age, residual confounding cannot be excluded. Our cohort comprised critically ill children, and whether these findings extend to adults or to less severely ill children is unknown. Finally, classifiers were evaluated by cross-validation within a single cohort, and external validation will be necessary, though suitable cohorts with paired airway transcriptomic and proteomic data do not currently exist.

Translating these findings into clinical assays will require validation of classifiers in independent cohorts, measured on platforms suited to clinical deployment rather than discovery, and extension to the diagnostically uncertain patients in whom the clinical need may be greatest. Determining whether these signatures are recoverable from more accessible respiratory specimens, and whether they are reproducible in less severely ill and adult populations, will be equally important, and longitudinal sampling would further clarify how the transcriptome and proteome relate to one another over the course of infection.

Taken together, our findings demonstrate that both the lower airway transcriptome and proteome capture the core biological features of LRTI, despite low correlation between expression levels of most individual genes and their matched proteins. Both enable accurate, pathogen-agnostic diagnosis, but the biomarkers they favor and the patients they identify differ. The choice of modality is therefore substantive rather than technical. This has implications beyond diagnosis: transcriptomic evidence is widely used to nominate therapeutic targets, yet the druggable entity is the protein, and our data indicate that transcript abundance is an unreliable proxy for it in the lower airway. The same caution applies to prognostic enrichment and pharmacodynamic endpoints, which increasingly rely on single-modality host profiling. More broadly, paired transcriptomic and proteomic profiling of tracheal aspirates from critically ill children provides a gene-protein resource for the lower airway, which is publicly available to facilitate further investigation.

## METHODS

### Study cohort and ethics approval

This study analyzes a prospective cohort of mechanically ventilated children admitted to eight US pediatric ICUs in the Eunice Kennedy Shriver National Institute of Child Health and Human Development Collaborative Pediatric Critical Care Research Network (CPCCRN) between February 2015 and December 2017.^44^ Patients analyzed here were a subset of those previously reported (**Figure S1**).^23^ To be eligible for this cohort, children were aged 31 days-18 years, expected to require mechanical ventilation via endotracheal tube for at least 72 hours, and had a TA sample collected <24 hours after intubation. Exclusion criteria included prior study enrollment, presence of a tracheostomy tube, prior mechanical ventilation during the hospitalization, and limitations of care. Informed consent was obtained from parents or guardians, and a waiver of consent permitted TA collection from standard-of-care endotracheal suctioning until parents or guardians could be approached. The cohort study was approved by the University of Utah central IRB #00088656.

### Clinical adjudication of LRTI status

LRTI status was adjudicated retrospectively, as previously described.^23^ At least two study-site physicians with expertise in pediatric critical care and/or infectious diseases independently reviewed all clinical, laboratory, microbiologic, and radiographic data available through the end of hospitalization. Patients were assigned to one of four categories based on the combination of clinical diagnosis and standard-of-care microbiologic testing (nasopharyngeal multiplex viral PCR and/or semiquantitative bacterial culture of tracheal aspirate, sputum, or bronchoalveolar lavage fluid), incorporating the CDC/NHSN PNU1 criteria^45^: Definite LRTI (clinical diagnosis with supportive microbiology), Suspected LRTI (clinical diagnosis without microbiologic findings), Indeterminate (microbiologic findings without clinical suspicion of LRTI), and No Evidence of LRTI (a clear noninfectious cause of respiratory failure with no microbiologic evidence of infection).

Only patients adjudicated as Definite LRTI or No Evidence of LRTI were analyzed here, referred to throughout as LRTI and controls.

### Specimen collection and processing

TA was collected <24 hours after intubation, and specimens were divided for transcriptomic and proteomic profiling. For RNA sequencing, whole TA was mixed equivolume with DNA/RNA Shield (Zymo Research) and stored at -80°C. For proteomics, TA was centrifuged at 15,000×g for five minutes at 4°C, and the supernatant was frozen at -80°C <30 minutes after collection.

### RNA sequencing and host gene count generation

Host transcriptomic data were generated as part of a previously published study.^23^ RNA was extracted from 300μL of TA using bead-based lysis and the AllPrep DNA/RNA kit (Qiagen), including a DNase treatment step. Sequencing libraries were prepared using the NEBNext Ultra II Library Prep Kit (New England BioLabs), and human ribosomal RNA was depleted prior to library amplification using the Cas9-based Depletion of Abundant Sequences by Hybridization (DASH) method.^82^ Libraries underwent 150bp paired-end sequencing on an Illumina NovaSeq 6000. Following demultiplexing, reads were pseudo-aligned with kallisto,^83^ including bias correction, to an index of all transcripts from human protein-coding and long noncoding genes (Ensembl v99). Gene-level counts were generated from transcript-level abundance estimates using the R package tximport with the scaledTPM method.^84^

### SomaScan proteomics

Protein abundance was measured from TA supernatant using the SomaScan 7k aptamer-based assay (SomaLogic).^85,86^ Normalized relative fluorescence units were log-transformed prior to analysis. Aptamers were annotated to gene symbols, UniProt identifiers, and gene names using the R package SomaScan.db. Aptamers without a gene symbol annotation were excluded, and for proteins targeted by more than one aptamer, values were averaged across aptamers to yield a single measurement per protein. To enable comparison between modalities, transcriptomic and proteomic data were matched on gene symbol.

### Respiratory pathogen burden by metatranscriptomics

Respiratory pathogens were identified from TA RNA sequencing data as previously described.^22,23^ Taxonomic alignment of non-host reads was performed using the CZID metagenomic pipeline, and background contaminants were identified and removed using a negative binomial model fit to water controls processed alongside patient specimens.^87^ Known pathogenic respiratory viruses were considered present at an abundance exceeding 0.1 RPM. Known pathogenic respiratory bacteria were identified using a previously established rules-based algorithm^22,23^ that identifies taxa disproportionately dominant relative to the surrounding lung microbiome. For each patient, pathogenic viral and bacterial abundance were quantified as the summed RPM of the pathogens identified in each category.

### Differential expression/abundance and pathway analysis

Differential expression (RNA) and differential abundance analyses (protein) were performed with the R package limma, comparing LRTI patients with controls and adjusting for age and sex as covariates.^88^ For transcriptomic data, genes were retained if they had >10 counts in >20% of samples, counts were normalized using the trimmed mean of M-values (TMM) method in edgeR, and voom transformation was applied prior to linear modeling.^89^ For proteomic data, linear models were fit directly to log-transformed normalized protein abundance. In both cases, gene- and protein-wise linear models were fit, and moderated t statistics were computed by empirical Bayes moderation. P values were adjusted for multiple testing using the Benjamini-Hochberg method, and an adjusted P<0.05 was considered significant. Gene set enrichment analysis was performed separately on the transcriptomic and proteomic results using the R package clusterProfiler.

Analytes were ranked by log_2_ fold change, and enrichment was tested against the MSigDB Hallmark and Reactome collections, obtained using the R package msigdbr.^90^ Normalized enrichment scores and Benjamini-Hochberg adjusted P values were computed for each gene set, with an adjusted P<0.05 considered significant. For correlations with pathways of interest (Hallmark “interferon alpha response” and Reactome “neutrophil degranulation”), pathway scores were computed for each patient as the mean of z-scored values across analytes in the gene set, calculated separately for transcriptomic and proteomic data.

### Classifier development

Diagnostic classifiers were developed with stratified 5-fold cross-validation, using identical fold assignments for the transcriptomic and proteomic workflows. All preprocessing and feature selection were performed within training folds only to avoid data leakage. For transcriptomic data, variance-stabilizing transformation was applied using DESeq2 in the training fold and then transferred to the held-out fold.^91^ Proteomic data were analyzed as log-transformed normalized relative fluorescence units. Features were selected by LASSO logistic regression using glmnet, with an inner 5-fold cross-validation within each training set to select the regularization parameter.^92^ Features were taken at two levels of sparsity: lambda.1se and the largest lambda that yielded exactly three nonzero coefficients. Single analyte classifiers were built using FABP4 and ISG15 alone. For each feature set, a random forest of 10,000 trees was fit to the training samples using the R package randomForest and applied to the held-out fold to generate an out-of-fold probability of LRTI for each patient, yielding a single prediction per patient for each classifier.^93^ Performance was assessed by AUC, computed from pooled predictions using the R package pROC, and performance of classifiers were compared using DeLong’s test.^94^

### Statistics

Analyses were performed in R (version 4.6.1). Patient characteristics were compared between LRTI patients and controls using Wilcoxon rank-sum tests for continuous variables and Fisher’s exact tests for categorical variables. Correlations between transcriptomic and proteomic measurements were assessed using Pearson correlation on z-scored gene and protein measurements; relationships between pathogen abundance and host response were similarly evaluated by Pearson correlation of standardized expression or pathway score against log_10_ transformed abundance. To test whether these relationships differed by either modality or infection status, linear models including interaction terms were fit. P values from analyses involving multiple comparisons were adjusted using the Benjamini-Hochberg method.

### Data and code availability

Host gene counts and microbial taxon counts for this cohort were generated in a previously published study^23^ and are available in the NCBI Gene Expression Omnibus under accession GSE212532 and at https://github.com/eranmick/pediatric-mNGS-LRTI-classifier. Deidentified clinical metadata, proteomic measurements, and the host gene counts and background-filtered microbial reports subset to the 105 patients analyzed here, together with all code required to reproduce the analyses presented in this manuscript and its supplement, are available at https://github.com/infectiousdisease-langelier-lab/RNA-Prot-LRTI-Classifiers-clean. Patients are identified by study-specific sample identifiers consistent across all data files.

## Data Availability

Host gene counts and microbial taxon counts for this cohort were generated in a previously published study23 and are available in the NCBI Gene Expression Omnibus under accession GSE212532 and at https://github.com/eranmick/pediatric-mNGS-LRTI-classifier. Deidentified clinical metadata, proteomic measurements, and the host gene counts and background-filtered microbial reports subset to the 105 patients analyzed here, together with all code required to reproduce the analyses presented in this manuscript and its supplement, are available at https://github.com/infectiousdisease-langelier-lab/RNA-Prot-LRTI-Classifiers-clean. Patients are identified by study-specific sample identifiers consistent across all data files.

## ACKNOWLEDGEMENTS

We thank all patients and families for contributing to this study. We acknowledge the contributions of the investigators and research coordinators at each of the study sites. This study was supported by the Collaborative Pediatric Critical Care Research Network, the Eunice Kennedy Shriver National Institute of Child Health and Human Development, the National Heart, Lung and Blood Institute UG1HD083171 (PMM), 1R01HL124103 (PMM), R01AI182308 (PMM and CRL), 5R01HL155418 (CRL), the Chan Zuckerberg Biohub (CRL), and Arkansas Children’s Research Institute (PMM). The study sponsors were not involved in study design, in the collection, analysis, or interpretation of data, in writing of the manuscript, or in the decision to submit the manuscript for publication.

