## Supplementary figures and tables for "Distinct pulmonary transcriptomic and proteomic host signatures enable accurate diagnosis of severe lower respiratory tract infection"

1. SUPPLEMENTARY FIGURES

2. SUPPLEMENTARY TABLES

3. BREATH CONSORTIUM

**SUPPLEMENTARY FIGURES**

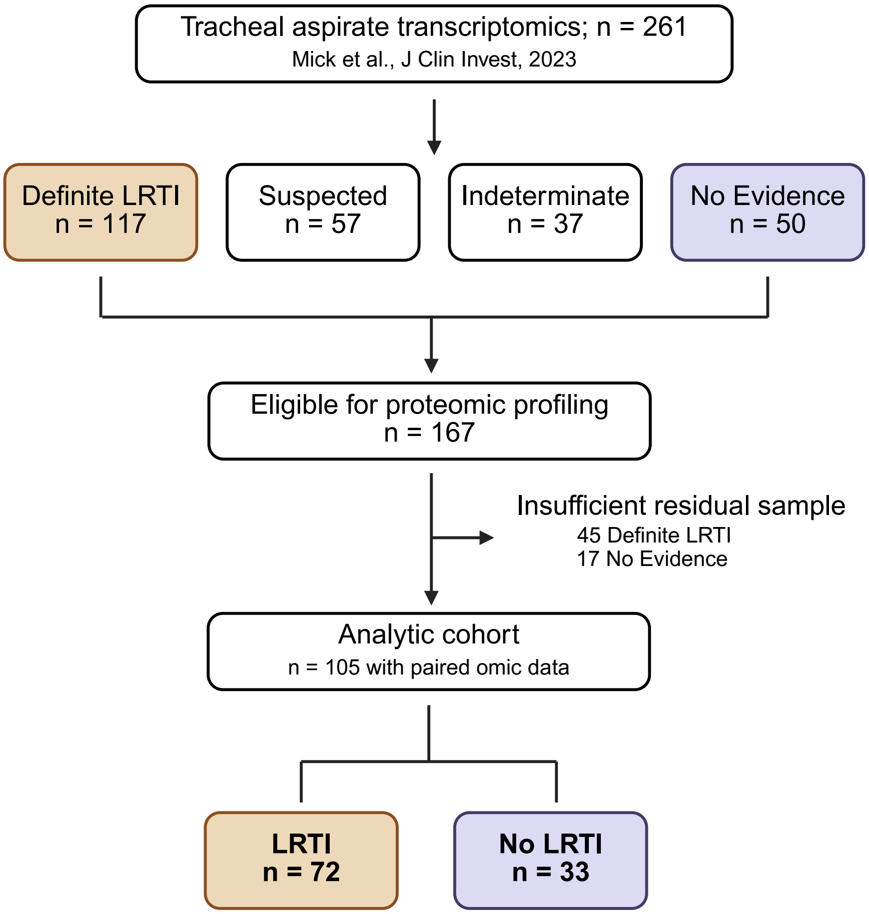


**Figure S1. Study flow diagram.** Patients were drawn from a previously described cohort of critically ill children with acute respiratory failure in whom lower airway transcriptomic sequencing was performed (n = 261). LRTI status was adjudicated retrospectively into four categories based on clinical diagnosis and standard-of-care microbiologic testing. Patients in the Definite LRTI and No Evidence groups with sufficient residual tracheal aspirate underwent proteomic profiling; 105 had paired transcriptomic and proteomic data passing quality control. Abbreviations: LRTI, lower respiratory tract infection.

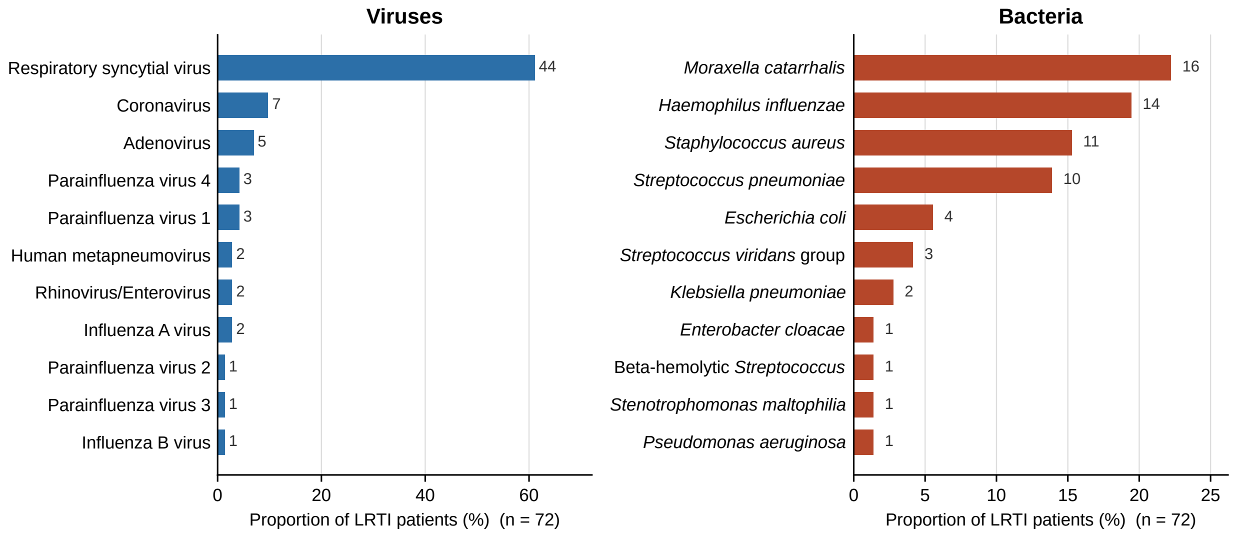


**Figure S2. Pathogens detected by clinical microbiologic testing in patients with lower respiratory tract infection (LRTI).** Proportion of patients in the LRTI group (n = 72) with each viral (left) and bacterial (right) pathogen identified by conventional clinical microbiologic testing at the treating institution. Numbers beside each bar indicate the number of patients. Proportions sum to more than 100% because 40 patients (56%) had codetection of more than one pathogen. Note the different x-axis scales for the two panels.

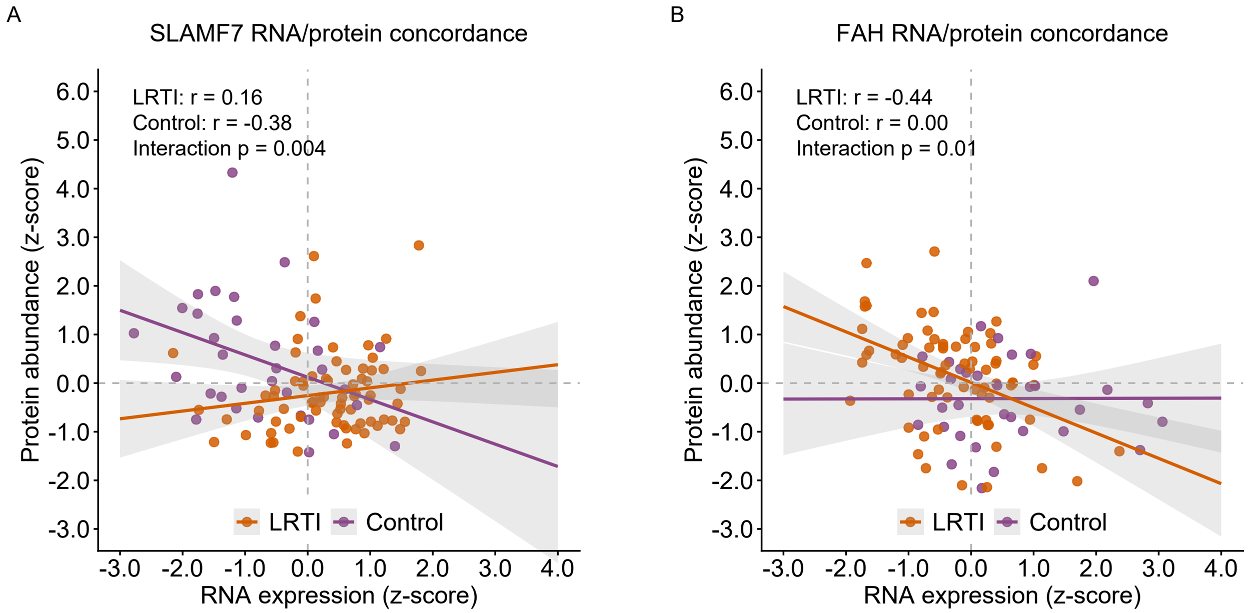


**Figure S3.** Scatterplots of z-scored protein abundance against z-scored RNA expression for SLAMF7 (A) and FAH (B), stratified by LRTI status. Points represent individual patients, lines indicate linear model fits, and shaded bands indicate 95% confidence intervals. Within-group Pearson correlation coefficients are annotated, along with the P value for the interaction term from a linear model testing whether the slope differed between groups. LRTI, lower respiratory tract infection.

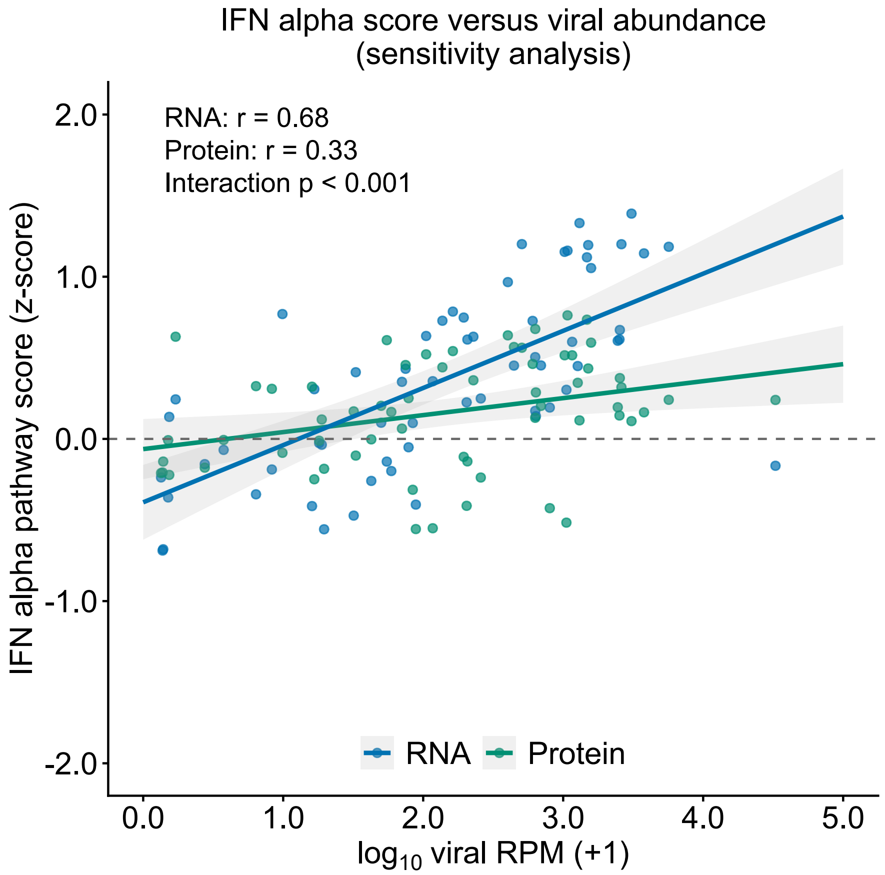


**Figure S4: The stronger association between viral abundance and the interferon-α response in the transcriptome is not explained by differences in analyte coverage between platforms.** Sensitivity analysis of Figure 3F, restricted to the 49 Hallmark "interferon alpha response" pathway analytes measured on both the transcriptomic and proteomic platforms. Composite interferon-α pathway scores were recalculated for the transcriptome (blue) and proteome (green) as the mean of z-scored values across this shared analyte set, and plotted against $log_{10}$-transformed summed viral abundance (reads-per-million, RPM) among LRTI patients with a respiratory virus detected by metatranscriptomic sequencing (n=62). Lines indicate linear model fits with 95% confidence intervals. Pearson correlation coefficients and the P value for the modality-by-viral-abundance interaction term are annotated. LRTI, lower respiratory tract infection; RPM, reads per million.

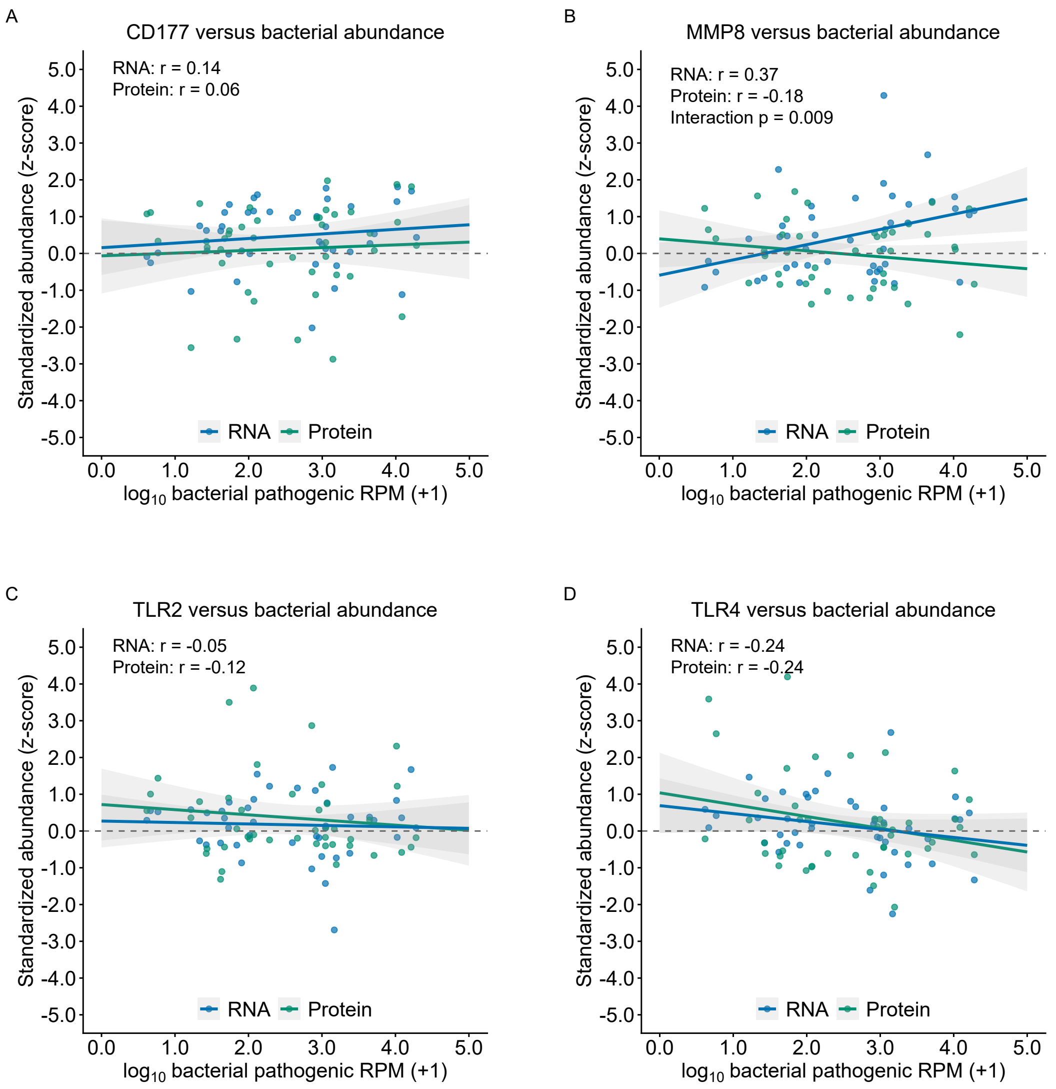


**Figure S5: Canonical antibacterial response analytes show no consistent association with bacterial abundance in either modality.** Standardized (z-scored) RNA expression (blue) and protein abundance (green) for CD177 (A), MMP8 (B), TLR2 (C), and TLR4 (D), plotted against $\log_{10}$-transformed summed pathogenic bacterial abundance (reads-per-million, RPM) among LRTI patients with a bacterial pathogen detected by metatranscriptomic sequencing (n=43). Points represent individual patients, lines indicate linear model fits, and shaded bands indicate 95% confidence intervals. Pearson correlation coefficients are annotated for each modality, along with the P value for the modality-by-bacterial-abundance interaction term where significant. LRTI, lower respiratory tract infection; RPM, reads per million.

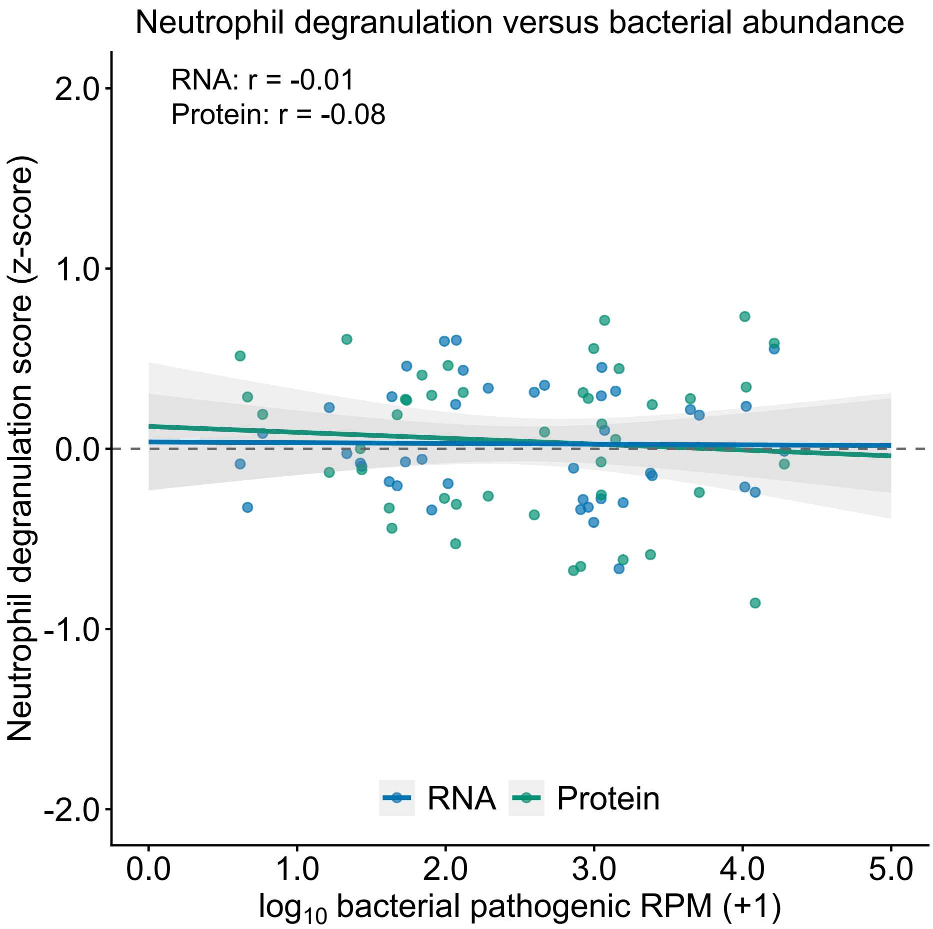


**Figure S6. The neutrophil degranulation response shows no association with bacterial abundance in either modality.** Composite Reactome "neutrophil degranulation" score, calculated separately for the transcriptome (blue) and proteome (green) as the mean of z-scored values across analytes in the gene set, plotted against $\log_{10}$-transformed summed pathogenic bacterial abundance (reads-per-million, RPM) among LRTI patients with a bacterial pathogen detected by metatranscriptomic sequencing (n=43). Lines indicate linear model fits with 95% confidence intervals. Pearson correlation coefficients are annotated for each modality; the modality-by-bacterial-abundance interaction term was not significant. LRTI, lower respiratory tract infection; RPM, reads-per-million.

### TABLES

| Symbol | Probe ID | UniProt | Fold 1 | Fold 2 | Fold 3 | Fold 4 | Fold 5 |
|---|---|---|---|---|---|---|---|
| ALDH3B1 | 12940-35 | A3FMP9 | | ✓ | | | |
| COL1A1 | 11140-56 | O76045 | | ✓ | | | |
| ENTPD1 | 3182-38 | P49961 | | ✓ | | | |
| FABP4 | 15386-7 | E7DVW4 | ✓ | ✓ | ✓ | ✓ | ✓ |
| GCH1 | 11185-145 | P30793 | ✓ | ✓ | ✓ | ✓ | ✓ |
| GHRL | 23581-131 | Q9UBU3 | | | ✓ | | |
| GNLY | 14102-6 | B4E3H9 | | ✓ | ✓ | ✓ | ✓ |
| INHA | 25921-3 | A8K8H5 | | ✓ | | | |
| ISG15 | 14148-2 | A0A096LPJ4 | ✓ | ✓ | | ✓ | |
| MCF2L | 13934-3 | H7C275 | | ✓ | ✓ | | ✓ |
| PCOLCE2 | 6081-52 | B2RCH9 | | | | | ✓ |
| QPCTL | 8866-53 | Q9NXS2 | | | ✓ | | ✓ |
| SPOCK3 | 9906-21 | B4DFW5 | | ✓ | | | |
| TIFA | 21113-3 | Q96CG3 | | ✓ | | | |
| TLL1 | 6383-90 | O43897 | | | ✓ | ✓ | |

**Table S1. Proteins selected by LASSO feature selection in the full proteomic classifier.** Proteins selected by LASSO logistic regression (lambda.1se) within each of the five cross-validation training folds. Fifteen unique proteins were selected in at least one fold. For each protein, the table lists the symbol, SomaScan SOMAmer reagent identifier (Probe ID), and UniProt accession. Checkmarks indicate the folds in which the protein was selected. Proteins are listed alphabetically by gene symbol.

| Symbol | Gene | Fold 1 | Fold 2 | Fold 3 | Fold 4 | Fold 5 |
|---|---|---|---|---|---|---|
| AC006055.1 | ENSG00000235886 | | | | ✓ | |
| AC007014.2 | ENSG00000274038 | | | | | ✓ |
| AC010255.2 | ENSG00000249610 | | ✓ | | | |
| AC013457.1 | ENSG00000259094 | | | | ✓ | |
| AC048344.4 | ENSG00000277342 | | | | | ✓ |
| AL161740.1 | ENSG00000236341 | ✓ | | | | |
| AP001453.2 | ENSG00000256116 | | | | | ✓ |
| ATP2C2 | ENSG00000064270 | | | | ✓ | |
| ATP2C2-AS1 | ENSG00000261286 | | ✓ | | | |
| CCNA1 | ENSG00000133101 | ✓ | | | | |
| CD69 | ENSG00000110848 | ✓ | | | | ✓ |
| CISH | ENSG00000114737 | | | | ✓ | |
| CHST3 | ENSG00000122863 | ✓ | ✓ | | | |
| COL21A1 | ENSG00000124749 | ✓ | | | | |
| CXCL5 | ENSG00000163735 | | ✓ | | ✓ | |
| DMTF1 | ENSG00000135164 | | ✓ | | | |
| FABP4 | ENSG00000170323 | ✓ | ✓ | ✓ | ✓ | ✓ |
| FCER1A | ENSG00000179639 | | ✓ | | | |
| FFAR3 | ENSG00000185897 | ✓ | ✓ | ✓ | ✓ | ✓ |
| F11R | ENSG00000158769 | | ✓ | | | |
| GBP5 | ENSG00000154451 | ✓ | | | | |
| GNLY | ENSG00000115523 | | ✓ | | ✓ | ✓ |
| IFITM1 | ENSG00000185885 | | ✓ | | | |
| IGFBP5 | ENSG00000115461 | | | | | ✓ |
| IRF7 | ENSG00000185507 | ✓ | | ✓ | ✓ | ✓ |
| ISG15 | ENSG00000187608 | ✓ | ✓ | | ✓ | |
| LINC01108 | ENSG00000226673 | | ✓ | | | |
| LRRC17 | ENSG00000128606 | | | | ✓ | |
| PSMB8 | ENSG00000204264 | | | | | ✓ |
| RFPL4AL1 | ENSG00000229292 | ✓ | | | | |
| SCGB1A1 | ENSG00000149021 | | | | | ✓ |
| SNURF | ENSG00000273173 | | | | ✓ | |
| SPATA20 | ENSG00000006282 | | | | | ✓ |

**Table S2. Genes selected by LASSO feature selection in the full transcriptomic classifier.** Genes selected by LASSO logistic regression (lambda.1se) within each of the five cross-validation training folds. Thirty-three unique genes were selected in at least one fold. For each gene, the table lists the gene symbol and the corresponding Ensembl gene identifier. Checkmarks indicate the folds in which the gene was selected. Genes are listed alphabetically by gene symbol.

| Symbol | Probe ID | UniProt | Fold 1 | Fold 2 | Fold 3 | Fold 4 | Fold 5 |
|---|---|---|---|---|---|---|---|
| FABP4 | 15386-7 | E7DVW4 | ✓ | | ✓ | ✓ | ✓ |
| GCH1 | 11185-145 | P30793 | ✓ | ✓ | ✓ | ✓ | ✓ |
| GNLY | 14102-6 | B4E3H9 | | ✓ | ✓ | | |
| ISG15 | 14148-2 | A0A096LPJ4 | ✓ | ✓ | | ✓ | |
| PCOLCE2 | 6081-52 | B2RCH9 | | | | | ✓ |

**Table S3. Proteins selected by LASSO feature selection in the parsimonious proteomic classifier.** Proteins selected by LASSO logistic regression constrained to three features per fold within each of the five cross-validation training folds. Five unique proteins were selected in at least one fold. For each protein, the table lists the gene symbol, SomaScan SOMAmer reagent identifier (Probe ID), and UniProt accession. Checkmarks indicate the folds in which the protein was selected; exactly three proteins were selected per fold. Proteins are listed alphabetically by gene symbol.

| Symbol | Gene | Fold 1 | Fold 2 | Fold 3 | Fold 4 | Fold 5 |
|---|---|---|---|---|---|---|
| CXCL5 | ENSG00000163735 | | ✓ | | | |
| FABP4 | ENSG00000170323 | ✓ | | ✓ | ✓ | ✓ |
| FFAR3 | ENSG00000185897 | | | ✓ | ✓ | |
| IFITM1 | ENSG00000185885 | | ✓ | | | |
| IRF7 | ENSG00000185507 | ✓ | ✓ | ✓ | ✓ | ✓ |
| RFPL4AL1 | ENSG00000229292 | ✓ | | | | |
| SPATA20 | ENSG00000006282 | | | | | ✓ |

**Table S4. Genes selected by LASSO feature selection in the parsimonious transcriptomic classifier.** Genes selected by LASSO logistic regression constrained to three features per fold within each of the five cross-validation training folds. Seven unique genes were selected in at least one fold. For each gene, the table lists the gene symbol and the corresponding Ensembl gene identifier. Checkmarks indicate the folds in which the gene was selected; exactly three genes were selected per fold. Genes are listed alphabetically by gene symbol.

**BREATH Consortium Members**

Lilliam Ambroggio (University of Colorado and Children’s Hospital Colorado, Aurora, CO); Aaron Bodansky (University of California, San Francisco, CA); Erin Carlton (University of Michigan, Ann Arbor, MI); Abagail Carolan (University of Michigan, Ann Arbor, MI); Gabrielle Cheney (Children’s Hospital Colorado, Aurora, CO); Natalie Cvijanovich (UCSF Benioff Children’s Hospital Oakland, Oakland, CA); Lanae Dailey (Children’s Hospital Colorado, Aurora, CO); Kevin J. Downes (Children’s Hospital of Philadelphia and Perelman School of Medicine, University of Pennsylvania, Philadelphia, PA); Lauren Gunn-Sandell (University of Colorado, Aurora, CO); CJ Jayachandran (University of Michigan, Ann Arbor, MI); Joshua Kennedy (University of Arkansas for Medical Sciences and Arkansas Children’s Research Institute, Little Rock, AR); Manisha Khakoo (University of California, San Francisco, CA); Teresa Kortz (University of California, San Francisco, CA); Charles R. Langelier (University of California, San Francisco, CA); Matthew Leroue (University of Colorado and Children’s Hospital Colorado, Aurora, CO); Ashlyn Madding (Arkansas Children’s, Little Rock, AR); Aline B. Maddux (University of Colorado and Children’s Hospital Colorado, Aurora, CO); Kelli McDonough (University of Michigan, Ann Arbor, MI); Erin McDowell (Children’s Hospital of Philadelphia, Philadelphia, PA); Patrick McQuillen (University of California, San Francisco, CA); Peter M. Mourani (University of Michigan, Ann Arbor, MI); Christina M. Osborne (Children’s Hospital of Philadelphia, Philadelphia, PA); Emma Peveri (University of California, San Francisco, CA); Ronald C. Sanders (University of Arkansas for Medical Sciences and Arkansas Children’s, Little Rock, AR); Anna Sharova (Children’s Hospital of Philadelphia, Philadelphia, PA); Marc Sheetz (Midwestern University, Downers Grove, IL); Kasturi Sinha (University of California, San Francisco, CA); Careese Thompson (Children’s Hospital Colorado, Aurora, CO); Katie Tolkacheva (Children’s Hospital of Philadelphia, Philadelphia, PA); Daniela Wild (University of California, San Francisco, CA); Masson Yates (Arkansas Children’s, Little Rock, AR); Nadir

Yehya (Children’s Hospital of Philadelphia and Perelman School of Medicine, University of Pennsylvania, Philadelphia, PA); Ming Ying Yeoh (University of California, San Francisco, CA); Matthew Zinter (University of California, San Francisco, CA).
